# Protective immunity to clinical malaria is modified by the genetic diversity of *P. falciparum* antigens

**DOI:** 10.64898/2026.01.30.26345157

**Authors:** Myo T. Naung, Ramin Mazhari, Rhea J Longley, Somya Mehra, Wilson Wong, Paolo Bareng, Moses Laman, Benson Kiniboro, Maria Ome-Kaius, Pascal Michon, James Beeson, Eizo Takashima, Hikaru Nagaoka, Shannon Takala-Harrison, Takafumi Tsuboi, Leanne J. Robinson, Ivo Mueller, Alyssa E. Barry

## Abstract

Prioritising malaria vaccine targets requires understanding immunity to genetically and structurally diverse parasite antigens, influencing antibody measurements and durability. We measured total IgG levels to 25 *Plasmodium falciparum* antigens and assessed their association with protection and antigen features. Antibodies were quantified in two longitudinal cohorts of Papua New Guinean children (5–14 years; n=647) experiencing high or moderate transmission. Associations between antibody levels and time to first clinical malaria episode were evaluated using Cox regression and Bayesian antibody-kinetics models, incorporating antigen genetic diversity and structural properties. In high-transmission settings, antibody levels were elevated and stable, with the strongest protection observed for conserved, low-diversity antigens dominated by the 3D7 variant and enriched for intrinsically disordered and alpha-helical regions. In moderate transmission, antibody levels were variable, decayed over time, and reflected recent exposure. These findings identify antigen diversity as a key modifier of malaria immunity and underscore the importance of antigen features.

## Introduction

Malaria remains a significant global health challenge, especially in sub-Saharan Africa, where around 95% of malaria cases and deaths occur^1^. In 2023, approximately 263 million people were infected with malaria worldwide, leading to over 597,000 deaths. Children under five are particularly vulnerable, accounting for about 80% of all malaria deaths in Africa^1^. The development and introduction of vaccines have been major milestones in the fight against malaria. Recently, two licensed malaria vaccines, RTS,S/AS01 (also known as Mosquirix) and R21/Matrix-M^2,3^, have been introduced in Africa. Both vaccines target the circumsporozoite protein (CSP) of the malaria parasite, *Plasmodium falciparum*, which is responsible for the deadliest form of malaria. While the introduction of RTS,S and R21 represents a significant breakthrough, both vaccines offer only moderate efficacy—estimated at 20–40%^2,3^—and further improvements are needed to meet the Malaria Vaccine Roadmap goal of achieving greater than 75% efficacy by 2030^4^. The search for an effective malaria vaccine remains challenging due to the extensive genetic and unique structural characteristics of *Plasmodium* parasite antigens^5,6^. Most malaria vaccines in development are based on subunit proteins using variants from a single strain^7^, but immunity is often strain-specific due to allele-specific immunity, and vaccine efficacy is greater against vaccine-like strains ^2,8^. One strategy is to include multiple variants of each candidate antigen to cover antigen diversity^4^. This approach could enhance vaccine efficacy by inducing immune protection against a greater proportion of strains. However, to realise this potential, an understanding of the extent to which genetic and structural diversity influence immunity, is needed.

Repeated exposure to infection is essential for the development of protective immunity to malaria, which is non-sterilizing and manifests as a reduction in symptoms and numbers of malaria episodes with age^9–11^. This immunity functions by limiting high-density parasitemia and inhibiting blood-stage replication^12^ and is associated with acquisition of a repertoire of antibodies to diverse antigens^9^. Experiments conducted in the 1960s, in which purified IgG from highly exposed adults in The Gambia were administered to twelve children (aged 4 months to 2 years) hospitalized with malaria, demonstrated the critical role that antibodies play in protection^13^. Naturally acquired and vaccine-induced antibodies elicit protection through direct inhibition of parasite invasion of host cells, complement fixation and killing, and opsonic phagocytosis and cellular cytotoxicity^14–19^.

Several studies have demonstrated an association between antibody levels to specific antigens and protective immunity against malaria, defined as an increase in the time to first clinical episode in high versus low responders, using a longitudinal cohort study design^20–24^. These studies have measured antibodies against antigen variants derived from a single strain, usually 3D7, resulting in an association between antibodies to individual variants and malaria outcome, but do not consider that new infections may carry a different variant. Our recent analysis of a publicly available database of 2,661 *P. falciparum* genome sequences demonstrates the high diversity of several vaccine candidates, with the 3D7 variant rare in many countries, whilst others are relatively conserved^5^. This diversity may result in a low protective efficacy since the variant used to measure responses represents only a small fraction of the circulating variants. The global database of *P. falciparum* genomes provides essential data to evaluate whether associations between antibody responses and malaria risk are impacted by genetic diversity, as well as other structural features which may influence immune responses.

We aimed to measure antibody levels in association with protection, with reference to antigen genetic diversity and structural properties. Protection associated with antibody levels was assessed using samples from two longitudinal paediatric cohorts from Papua New Guinea (PNG), conducted during high (2006) and moderate transmission periods (2013, i.e. after intensive control efforts). These cohorts have well-characterised population and individual differences in exposure and disease ^25–28^. Antibody responses at the beginning and end of the cohort study were measured to the 3D7 variant of 25 *P. falciparum* vaccine candidates and key invasion ligands^22,29–31^. By comparing antibody levels and protective immunity in these different transmission contexts (e.g. repeated infection vs few infections) we were also able to explore antibody decay. We developed a model to estimate population-level antibody kinetics (boosting and decay) accounting for the genetic diversity of these antigens. The results highlight the importance of considering antigen diversity in antibody studies and provide critical information for the prioritisation and development of vaccine candidates.

## Results

### Structural and population genetic features of antigens

We first quantified population genetic diversity of antigens included in this study using data from the PNG population of MalariaGEN Pf7 dataset (n = 222) calculating nucleotide diversity, haplotype diversity, prevalence of the 3D7 variant, genetic distance to the 3D7 variant, and *Tajima’s D*. These metrics capture both the magnitude of sequence variation and departures from neutral evolution, enabling us to distinguish highly conserved antigens from those under balancing or directional selection. In parallel, we assessed key structural features—including intrinsically disordered regions (IDR) and alpha-helical content—to contextualise how structural flexibility and secondary structure may influence antigen variability and immune recognition.

Overall, the results show broadly defined categories of antigens based on population-genetic and structural characteristics. Several antigens, such as MSP3, MSP4, MSP7, RALP1, EBA175 region III–V and TRAMP, were completely conserved in the PNG population, with no measurable genetic diversity and near-complete fixation of the 3D7 variant (≥90%). These conserved antigens tended to exhibit moderate intrinsic disorder (median = 0.40) and intermediate alpha-helical content (median = 0.37) (Table 1). In contrast, antigens such as AMA1, DBLMSP, CLAG8, and TRAP displayed markedly high genetic diversity, low 3D7 variant prevalence, and strongly positive *Tajima’s D* values, consistent with balancing selection (Table 1). These more diverse antigens were characterised by lower disorder scores (median = 0.35) and reduced alpha-helical content (median = 0.25). Taken together, these results reveal a consistent relationship between structural properties and population-genetic signatures i.e. antigens with more ordered structural features generally accumulate diversity and exhibit signatures of balancing selection, whereas antigens with greater structural flexibility generally remain nearly invariant.

**Table 1.**
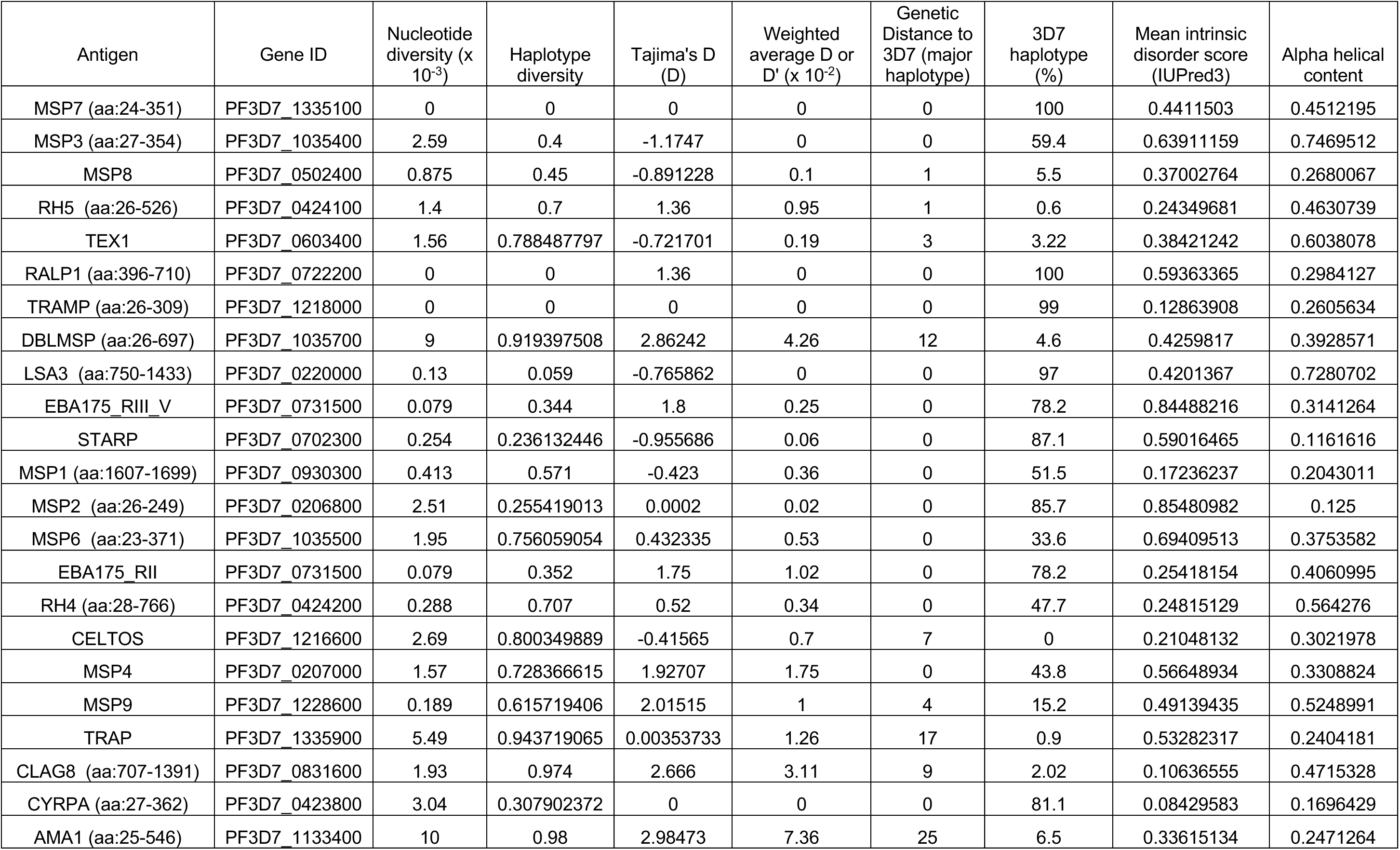
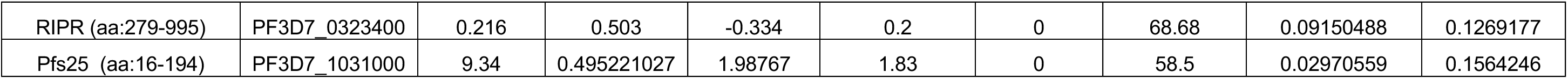
Summary results of population-genetic and structural measures for *P. falciparum* antigens.

### Antibody levels and dynamics

A total of 1,294 samples were screened for IgG levels using a multiplex bead array consisting of the 25 *P. falciparum* antigens including five full length, 20 ectodomain-removed or partially expressed antigen fragments, including two fragments from Erythrocyte Binding Antigen 175 (EBA175) (**Table S1 and S2**). Relative antibody unit (RAU) of IgG antibody levels and seroprevalence (proportion of samples with RAU greater than the mean RAU of negative controls + 3 standard deviation) varied among the 25 *P. falciparum* antigens tested (**Figure 1A, and S1**).

**Figure 1.**
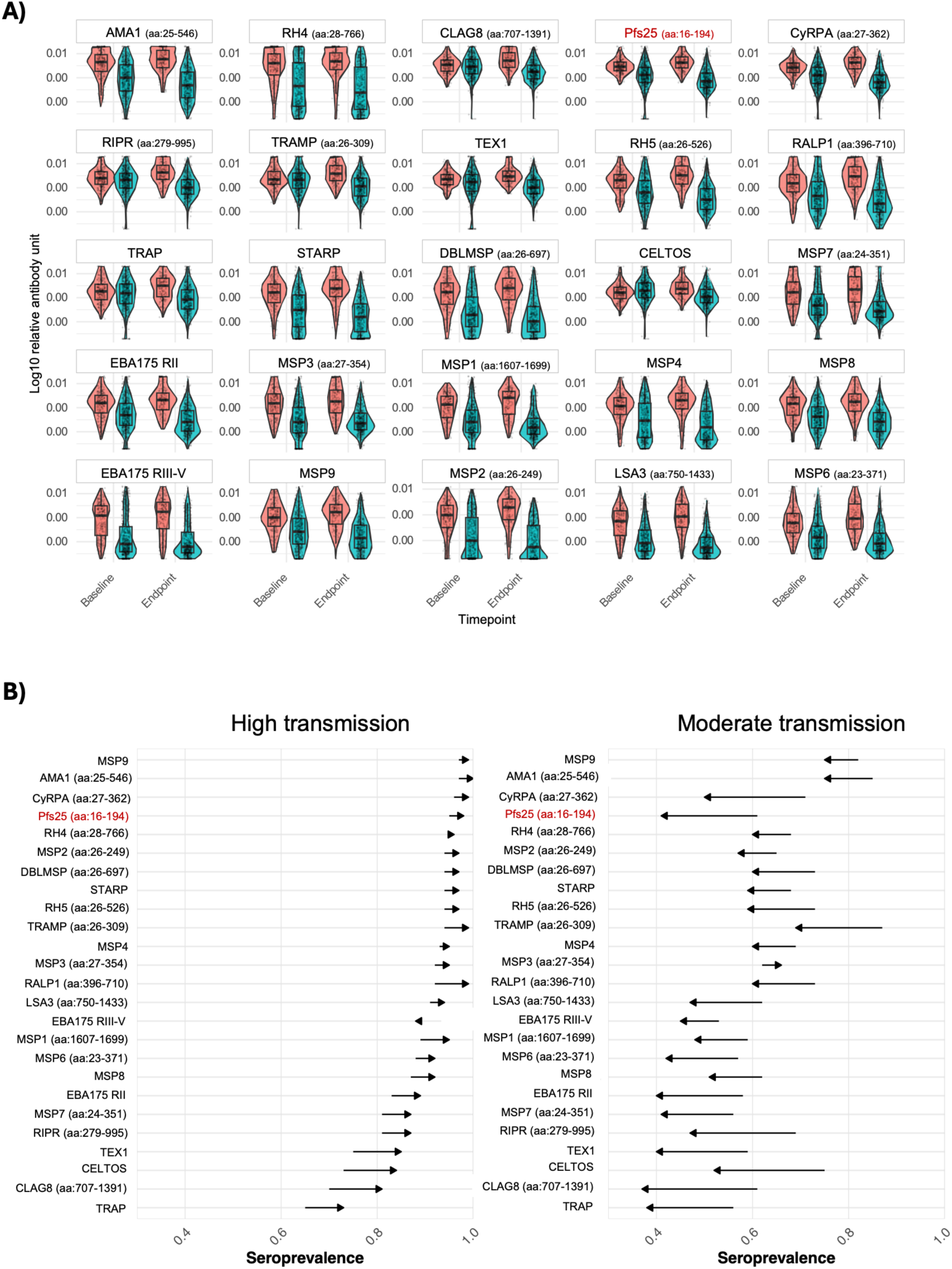
Maintenance of IgG antibody responses to 25 P. falciparum vaccine candidate antigens in high and moderate transmission cohorts from PNG. A) Distribution of log10 transformed RAU for each antigen for high transmission (red) and moderate transmission (green) cohort at both the first timepoint and endpoint. Antigens are ordered by median relative antibody units at high transmission cohort. B) Seroprevalence of antibodies to 25 P. falciparum proteins in Papua New Guinean children from different transmission settings highlighting how seroprevalence trends vary by cohort. For each antigen, the seroprevalence at two time points, the first timepoint and the study endpoint, is shown. The direction of the arrows indicates the change in seroprevalence from the first timepoint to endpoint, while the length of each arrow reflects the magnitude of that change. Antigens were ordered by their first timepoint seroprevalence within high transmission cohort.

For high transmission, antibody levels were significantly higher at the study endpoint compared to the first timepoint (*Mann-Whitney U test*, p < 0.05) except MPS3, MSP7, EBA175 RIII-V and RH4, which showed no increase (**Figure 1A**). Seroprevalence also increased with an average of 85% of individuals seropositive (range: 65-95%) at the first time point, and 90% (range: 81–100%) were seropositive at the endpoint (**Figure 1B**). The moderate transmission cohort exhibited the opposite pattern, where antibody levels were significantly higher at the first timepoint than the endpoint (*Mann-Whitney U test*, p < 0.05), for all antigens except MSP3 (**Figure 1A, and S1**). However, the patterns of antibody decline varied across antigens, with more pronounced differences between time points for TRAMP, CELTOS, CyRPA, CLAG8, Pfs25, RIPR, LSA3, MSP7, and TRAP. The average seroprevalence was 67% (range: 53-87%) at the first timepoint and only 52% (range: 40–75%) at the study endpoint across all tested antigens (**Figure 1B**). Notably, the highest seropositivity rates in both transmission settings were observed for AMA1, MSP9, DBLMSP and TRAMP (**Figure 1B**), consistent with their high immunogenicity^6,22,23,29^.

Interestingly, there were some correlations in antibody responses between antigens (Spearman correlation coefficient, *ρ* > 0.5) found in complexes or gene families, and these were consistent across different transmission settings (**Figure S2**). The strongest correlations were observed among antibodies targeting the RH5-CyRPA-RIPR (RCR) complex, TRAMP and the RCR complex, between CyRPA and Pfs25, between AMA1 and MSP-family antigens, and between EBA175 RII and EBA175 RIII-V. Importantly, none of these correlations exceeded a Spearman’s ρ of 0.7, indicating moderate—rather than strong—overlap in antibody recognition. No negative correlations were observed.

### Factors contributing to antibody decay

To investigate factors contributing to the observed variability in antibody decay, we performed a retrospective multivariate linear regression analysis using categorical antibody data from the final time point. This analysis was done only using the moderate transmission cohort as we wanted to understand antibody decay in the absence of natural infection (**Figure 2**). *P. falciparum* age and time to infection categories accounted for the largest proportion of explained antibody variance for most antigens. This was more pronounced for more conserved antigens such as MSP4, MSP7, TRAMP and RALP where the predominant circulating haplotype closely matches the assessed 3D7 strain^5^. In contrast, non-blood stage antigens such as TRAP (sporozoite stage), CELTOS (gametocyte and sporozoite stage), with high sequence diversity, and a low prevalence of 3D7 in natural populations^5^, may limit the ability to detect these associations. Thus, these antigens showed a low proportion of explained variance. Our results also reveal that *P. vivax* malaria infection has minimal impact on antibody variation confirming a lack of cross-species protection to the antigens studies.

**Figure 2.**
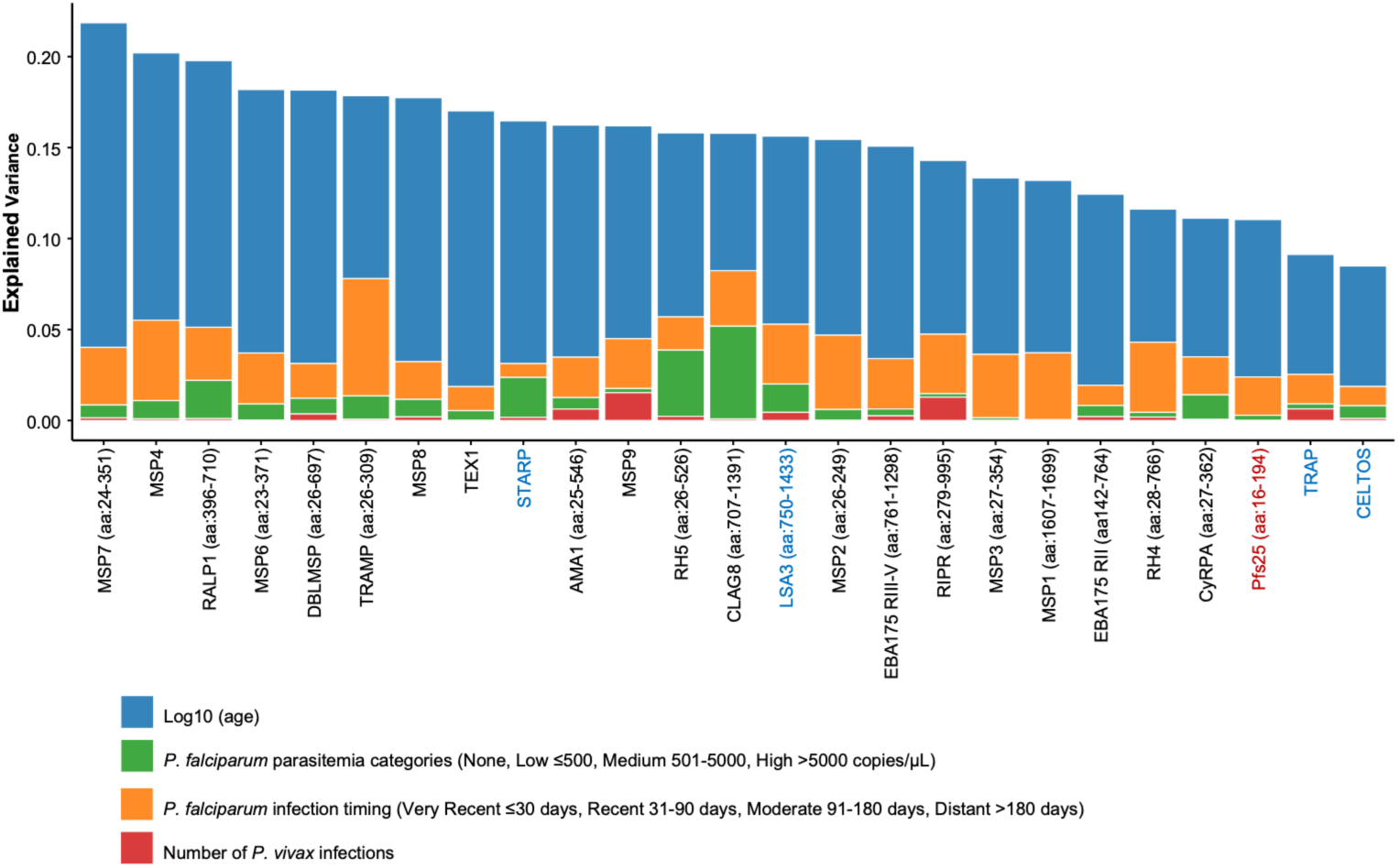
Contribution of variables to the variance in antibody responses using categorical modelling. Each bar represents the variance explained for a specific antigen using categorical variables that avoid collinearity between infection number and timing. The area of each coloured segment within a bar corresponds to the proportion of variance accounted for by known factors, ordered from left to right by: infection timing, infection frequency), log10-transformed age, and number of P. vivax infections. Antigens are ordered by total explained variance (highest to lowest). The colour of the x-axis labels indicates the life cycle stage of Plasmodium falciparum to which each antigen belongs: black for blood-stage, blue for pre-erythrocytic stage, and red for gametocyte-stage antigens. This categorical approach provides more reliable variance estimates compared to continuous variable models that suffer from collinearity between infection number and timing (r = -0.573, p < 0.001).

### Protective immunity

Building on these exposure-driven differences in antibody dynamics, we next investigated whether antibodies to each antigen were associated with protective immunity, based on the time to first malaria episode after the antibody measurements. In the high-transmission cohort, most *P. falciparum* antigens showed a statistically significant association between higher antibody levels and reduced risk, with adjusted hazard ratios (*aHR*) ranging from 0.21 [0.11–0.40] to 0.82 [0.50–1.35] (**Figure 3**). Conversely, in the moderate transmission cohort, a positive trend was observed between antibodies and risk of *P. falciparum* clinical malaria for most antigens, which is consistent with a lower level of immunity overall in this moderately exposed cohort^32^. The adjusted hazard ratios (*aHR*) for this association ranged from 0.88 [0.59–1.31] to 1.82 [1.17–2.73] (**Figure 3B**).

**Figure 3.**
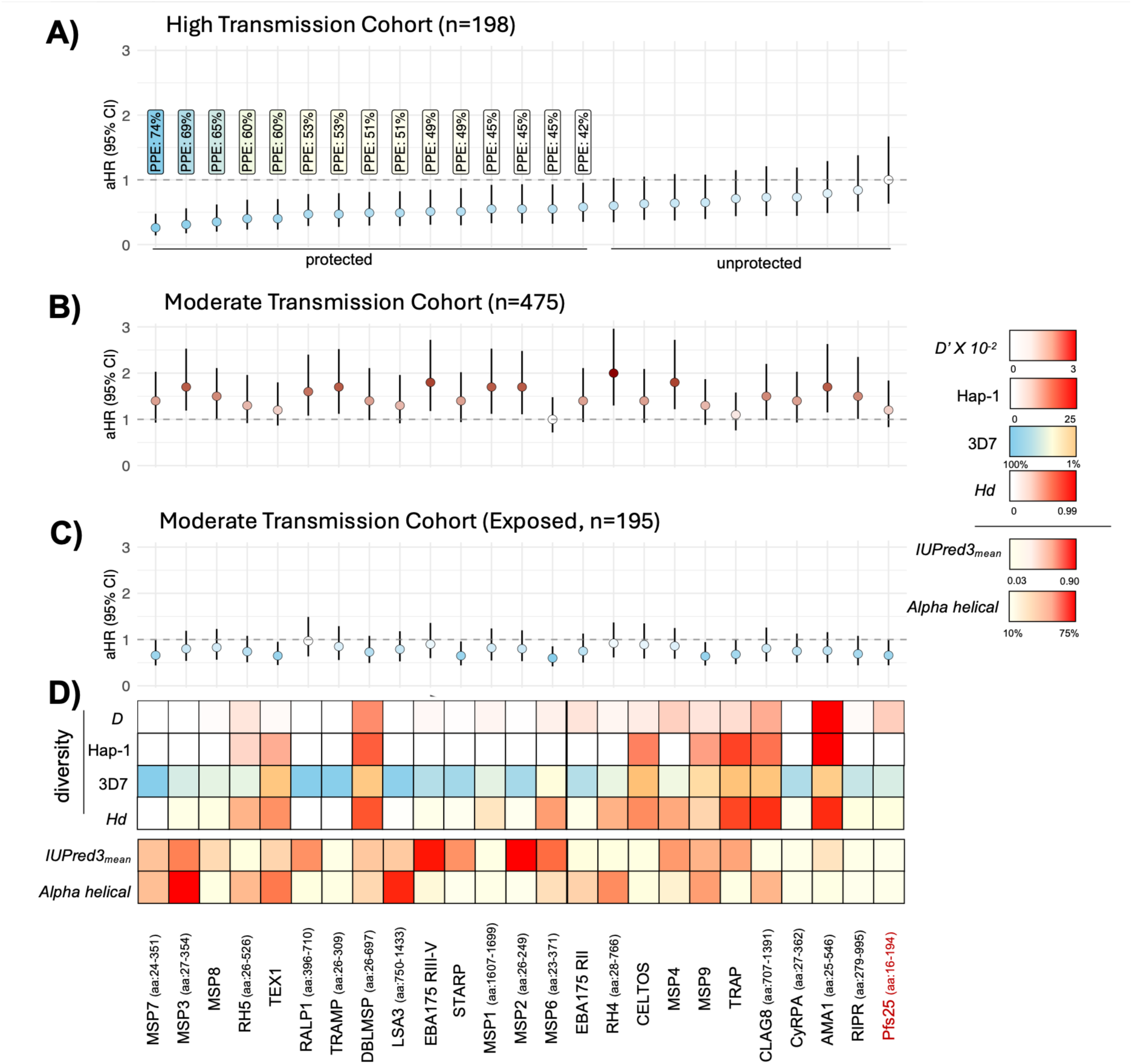
P. falciparum Antigen-Specific Protective Immunity in High and Moderate Transmission Settings (n = 196 for high transmission; n = 495 for moderate transmission) **A**) Adjusted Hazard Ratios (aHR) with 95% confidence intervals are shown. Each antigen is plotted on the x-axis (including their expressed regions), ranked from the lowest to highest aHR (MPS3 has the lowest, and Pfs25 has the highest). aHR values were calculated using a Cox regression model, assessing the risk of P. falciparum-specific clinical malaria (time to the first clinical P. falciparum infection episode). Error bars represent 95% confidence intervals, and the horizontal line at aHR = 1 indicates 0% protection. Potential Protective Efficacy (PPE) is represented as percentages, with 100% indicating complete theoretical protection against clinical malaria and 0% indicating no association with protection. Antigens with PPE > 50% were defined as having high PPE. PPE values were not calculated for antigens with insignificant aHRs. **B)** In the moderate transmission cohort, higher antibody levels were associated with an increased risk of malaria, likely reflecting recent exposure without adequate protective immunity. **C)** In the previously exposed subgroup of the moderate transmission cohort, some antigens exhibited protective associations similar to those in the high transmission cohort, though with lower efficacy. **D)** Diversity metrics and structural properties of each antigen are as follows: Tajima’s D (D) ranges from 0 (white) to 3 (red), while Hap-1 represents the genetic distance from 3D7 for the most common haplotype found in PNG, ranging from 0 (white) to 25 non-synonymous SNPs (red). The frequency of the 3D7 haplotype in PNG is represented as 100% (sky blue) to less than 1% (orange). Haplotype diversity (Hd) is shown on a red gradient, increasing from low (0) to high diversity (0.99). IUPred3mean indicates the mean disorder scores for each vaccine candidate antigen, visualized as a red gradient from low (0.03) to high (0.90). Finally, the proportion of alpha-helical residues for each vaccine candidate antigen is also represented as a red gradient, from low (10%) to high (75%). Gametocyte antigen, Pfs25 (assumed as negative control) is highlighted in red.

To confirm the role of exposure in the acquisition of protective immunity, individuals from the moderate transmission cohort were classified into two groups based on *P. falciparum* blood-stage infection during the follow-up period, as measured by qPCR: (i) exposed (at least one infection, n = 195) and (ii) low exposure (no infections, n = 280). While individual antigen-level associations differed, the overall pattern in this group resembled that of the high-transmission cohort, showing a trend toward protection against clinical malaria (**Figure 3C**). A significant association was observed for *P. falciparum* antigens including MSP6 and MSP7—most of which (except MSP9 and TRAP), were also involved in statistically significant protective responses in the high-transmission setting.

### Antigen genetic diversity and protective immunity

Antigens with the strongest associations with protection in the high transmission setting—such as the blood-stage antigens MSP3, MSP7, MSP8, RALP1, and TRAMP—were characterized by lower genetic diversity and balancing selection, as indicated by lower weighted average *D* or *D’* value (*Mann–Whitney U test*, p = 0.00082) (**Figure 3A, 3D; Table 1**). In addition, the dominant variant is either identical to, or similar to the 3D7 variant (**Figure 3A and D**, **Table 1**). In contrast, antigens such as AMA1, which exhibit high genetic diversity and strong balancing selection (high weighted average D or D′), showed only weak associations with protection (Figure 3A, 3D; Table 1). Notably, the antigens associated with protection amongst exposed individuals from moderate transmission settings—such as MSP6 and MSP7— showed a similar pattern, with generally lower weighted average D or D′ values (*Mann–Whitney U test*, p < 0.001), again indicating reduced balancing selection and greater 3D7-like haplotype prevalence (Table 1; Figure 3C, 3D). Pfs25, a gametocyte-stage antigen, showed the weakest association with protection given its role in the transmission, rather than blood stage. In addition, caution is needed for interpretation of LSA3 and MSP2 as the quality of sequencing data for these genes is compromised by stretches of tandem repeats that are difficult to assemble using short read sequencing^5,33^, and thus may result in an underestimation of their diversity.

### Structural properties and protective immunity

Antigens such as DBLMSP, MSP3, MSP4, MSP7, RALP1, EBA175 region III–V and TRAMP, exhibit a high proportion of intrinsically disordered residues (approximately 50% of total residues) and a significant presence of alpha-helical secondary structures, particularly evident in MSP3 and MSP7 (**Figure 3A and D**, **Table 1**). The low diversity, selectively neutral, antigens likely contribute to the association of antibodies with protective immune responses. On the other hand, antigens that showed no significant association with protection from clinical malaria in the high transmission setting including AMA1, CLAG8, and TRAP, were diverse with evidence of balancing selection, and had a lower proportion of intrinsically disordered regions and limited alpha-helical content (*Mann-Whitney U test*, p = 0.0231) (**Figure 3A and D**, **Table 1**).

### Antibody kinetics modelling

To formally measure the antibody kinetics associated with the prescribed antigen features, we adapted a previously validated Bayesian model^34^ by incorporating genetic diversity parameters for each antigen (**Table 1**). The model was designed to capture antigen-specific population-level patterns in the boosting and decay of antibody profiles, rather than estimating individual-level trajectories, while estimating the average antibody kinetics and their variance at the population level for the moderate transmission cohort. Antibody data from the moderate transmission cohort was used because it represents a setting with minimal recurrent exposure, allowing population-level antibody decay to be observed more clearly in the absence of ongoing infection. The model was applied independently to data for each antibody response, with or without adjustment for genetic diversity, using a Bayesian framework with mixed-effect methods (**Figure S3**).

Although precise individual antibody half-lives could not be estimated in this study due to the availability of only two timepoints, we observed comparative, population-level patterns in normalized antibody durability scores across antigens, with some evidence of association with protective immune responses observed in the high transmission setting. Antigens that showed strong associations with protection from clinical malaria—such as MSP8, DBLMSP, TEX1 and TRAMP—also exhibited higher antibody durability scores compared to other antigens (**Figure 3 and 4**). In contrast, AMA1, which corresponds to limited protection from clinical malaria in the high transmission setting with low 3D7 haplotype prevalence, showed the lowest antibody durability (**Figure 3 and 4**). TRAP, CELTOS, and STARP were excluded from this analysis due to high signal-to-noise ratios or inability of the model to reach convergence at the population level.

**Figure 4.**
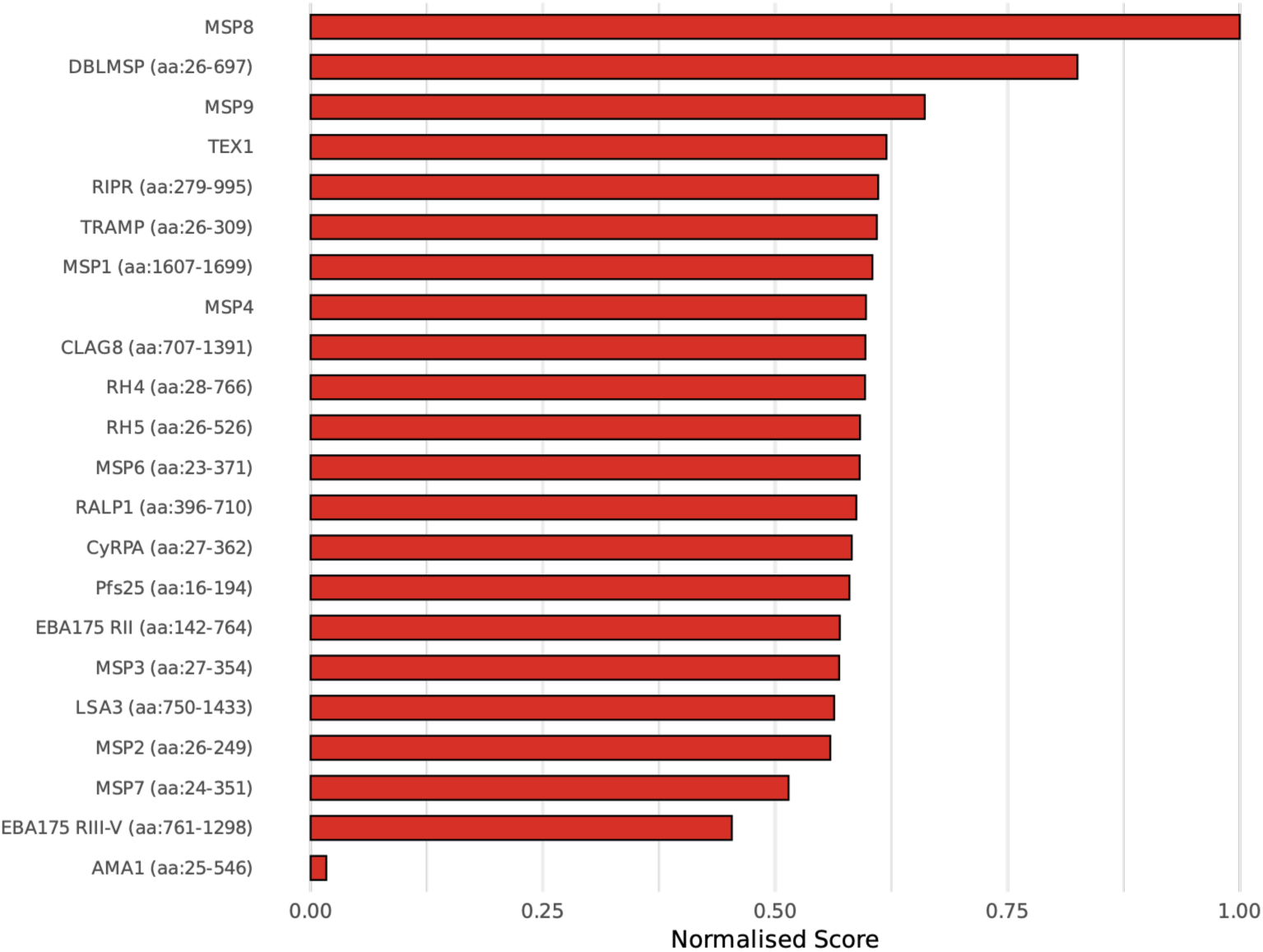
Composite normalized antibody durability scores for Plasmodium falciparum antigens. Each bar represents a composite score summarizing the modelled half-lives of antigen-specific antibody responses. For each kinetic component—short-lived, intermediate, and long-lived—half-life values were normalized across antigens, and then integrated using weighted contributions (20%, 20%, and 60%, respectively). Half-lives were then normalized from 0 (shortest) to 1 (longest). Antigens are ordered by their final composite scores, with higher values indicating relatively more durable antibody responses within the antigen panel.

## Discussion

The genetic diversity of parasite antigens is a major limitation for the development of effective and long-lasting malaria vaccines^2,5^. Only a few studies have evaluated antibodies to multiple antigens concurrently in the same population, and show mixed results^22,23,29,35–38^. The results may be influenced by malaria transmission intensity which could drive the rate of acquisition of antibodies to a wide repertoire of variants^35,39,40^, or above a protective titre^24,41^. By analysing antibody levels and dynamics in two cohorts, we found that the rate of acquisition of antibodies to different antigens varies, as do measures of protective immunity. Insights into antibody decay were revealed at different levels of malaria transmission which were modified by both antigen diversity and the frequency of exposure to circulating variants. By examining these patterns with reference to antigen diversity and structural properties, the investigation has revealed the importance of individual antigen characteristics in observed antibody responses and protection against malaria episodes.

Antibody dynamics were highly dependent on the transmission setting. At high transmission, sustained exposure throughout the follow-up period drives antibody acquisition and maintenance, resulting in a substantial increase in antibody magnitude and seroprevalence, with near universal seropositivity amongst the cohort participants by the study endpoint. In the moderate transmission setting however, antibody responses waned over time, with the rate depending on specific antigen features. By comparing these two exposure environments, we were able to contrast conditions of ongoing boosting versus limited exposure, demonstrating that continuous reinfection is required to develop and maintain protective immunity to clinical malaria^9,10,42–44^. The rise in antibody levels in the high-transmission cohort, alongside the consistent decline observed in the moderate-transmission cohort, clearly indicates that sustained exposure is necessary for boosting antibody titres and maintaining them above protective thresholds. This effect was more pronounced for conserved antigens such as MSP7, TRAMP where the majority of Papua New Guinean parasites carry the 3D7 variant tested in the antibody assays, and resulting in frequent antibody boosting at high transmission. Lower antigen diversity therefore contributes to the maintenance of antibodies over time (through boosting), while higher antigen diversity limits the detection of the full repertoire of antibodies present in a given individual, probably providing only the protective efficacy against infections carrying that particular variant. New infections carry distinct variants resulting in novel antibodies, that may not be detected—or detected less efficiently—in the 3D7 variant based assay, resulting in lower apparent antibody levels. Further studies of allele-specific responses at the individual level, specifically by analysing different variants of each antigen is an important next step, because population-level analyses may obscure important variation in exposure history, immune responsiveness, and antigen recognition.

Antibodies against most of the 25 antigens were associated with protection against clinical *P. falciparum* malaria in the high-transmission setting, but not in the moderate-transmission setting. This can be explained by lower immunity due to both an absence of boosting (conserved antigens) and gaps in the antibody repertoires (diverse antigens) of the latter cohort, and the continuous exposure required for acquisition and maintenance of antibodies above protective thresholds. This is again demonstrated by the close positive association between antibody levels and recent *P. falciparum* exposure in the moderate transmission setting, where people with higher antibody titres are at a higher risk of malaria exposure with clinical symptoms^23,32,45^. This cohort has limited immunity at the first timepoint and waning immune responses due to a lack of boosting during the follow-up period. Individuals with higher antibody levels, are at greater risk of developing symptomatic malaria than those with lower exposure because they have not yet acquired clinically immunity^24,46,47^.

Protective associations varied substantially among the 25 antigens. In general, antibodies targeting the more conserved antigens with predominantly 3D7 or 3D7-like variants, and limited balancing selection, were strongly associated with protective immunity especially within the high transmission setting. For instance, the protective associations were stronger for MSP3, MSP7, MSP8, and RALP1 which have low genetic diversity and variants identical to or similar to 3D7. Although some antigens were genetically diverse, antigens such as DBLMSP exhibited more durable antibody responses, possibly reflecting their immunogenicity, a limited number of antigenically distinct serotypes and a strong association with clinical immunity in the high transmission setting. Antibodies to MSP7 and RALP1 have been shown to be associated with protection against symptomatic malaria in previous studies^22,29^. On the other hand, antibody responses to diverse antigens with significant balancing selection such as CLAG8, and AMA1 with the exception of DBLMSP, were only weakly associated with protective immunity. Antibody responses against AMA1 and CLAG8 have lower antibody durability scores than MSP8, consistent with previous studies showing that allele-specific effects wane after six weeks ^6,48,49^. This suggests that exposure to conserved antigens in new infections would continually boost antibodies to the same epitopes, whereas for diverse antigens, new infections may stimulate the acquisition of antibodies to other variants. Genetic diversity, age, infection timing categories or *P. falciparum* infection frequency were major explanatory variables for the inter-individual differences in antigen-specific antibody responses. The antigen genetic diversity, and transmission setting, may therefore provide an explanation for previously reported mixed results regarding the protective associations of antibodies to *P. falciparum* antigens^35^. These results reinforce the role for repeated infection to induce protective immune responses, and this is particularly important for diverse antigens. For example, allele-specific protection has been demonstrated for antibodies to MSP2^23^ and the acquisition of highly functional antibodies to both major variants of MSP2 was found to be very strongly association with protection^29^.

Antigens that induced stronger protective immune responses, were also characterised by disordered regions. These disordered regions, also called intrinsically disordered regions (IDRs), are stretches of the protein that lack a fixed three-dimensional structure under physiological conditions. IDRs have previously demonstrated to have important molecular functions including receptor-ligand binding, DNA and RNA binding^50,51^ and are typically the targets of B-cell mediated immune mechanisms in malaria^52–55^. DBLMSP, MSP6, MSP7, TEX1, and STARP, which exhibited the strongest associations with protection in both transmission settings, are characterized by a high proportion of intrinsically disordered residues, a feature that may contribute to heightened immunogenicity and protective efficacy. A third (32.7%) of the *P. falciparum* proteome was predicted to be disordered including major surface antigens such as merozoite surface protein-2 (MSP2) and circumsporozoite protein (CSP)^53^. The CSP antigen included in the RTS, S/AS01 and R21/Matrix-M vaccine construct consists of an IDR with ‘NANP’ repeats targeted by antibodies with multiple functions ^8,18,56^. Our study supports a role for immunity specific to IDRs, and their protective efficacy against clinical disease. The functional relevance of these IDRs in malaria immunity likely extends beyond antigen stability. Their structural flexibility may enhance B-cell receptor recognition, facilitating stronger and more durable antibody responses^53^. Additionally, antigens with significant associations with protection also contained a higher proportion of alpha-helical regions such as in MSP3, MSP7 and RH5. These alpha-helices may complement disordered regions by providing structural motifs critical for host-parasite interactions and immune recognition. Alpha-helices and IDRs may define immunogenicity by promoting stable yet flexible antigen conformations that optimise B-cell receptor engagement and antibody binding^57^. This could represent another overlooked factor contributing to the variability in immune responses among the antigens contributing to their immunogenicity and ability to induce protective immune responses. However, robust functional studies are still needed to confirm and further elucidate these mechanisms.

The finding of high potential protective efficacy (60%) of RH5 at high transmission was notable, given it is in advanced clinical trials as a malaria vaccine^58^. The RH5-CYRPA-RIPR (RCR) complex is naturally immunogenic^22,59–62^, and we might be indirectly measuring the protective responses of the RCR complex. For instance, IgG responses to individual components such as RH5, CyRPA, or RIPR may in fact reflect cross-binding to epitopes presented at the interfaces between subunits, such as an RH5-specific IgG recognising an epitope at the RH5–CyRPA interface; however, none of these antigen–antigen correlations exceeded a Spearman’s correlation value of 0.7, indicating only moderate overlap in antibody recognition across the complex.

Our study has some limitations. Further insights in the rate of antibody decay associated with particular antigen features would be yielded by measuring antibody responses at additional time points between baseline and the end point of the follow-up period^34,63^. The temporal resolution with individuals contributing only two data points, limits the individual-level assessment of antibody kinetics, such as boosting, antigen-specific decay rates at each individual level, and mechanisms behind the maintenance of malaria-specific B-cell memory responses, potentially influencing the variability in individual immune responses and antibody kinetics. Additionally, other factors such as host genetics, lifestyle, and environmental variations may contribute to the unexplained variance observed in antibody decay profiles. Short read sequencing data, particularly for the *lsa3* and *msp2* genes were of low quality due to the presence of tandem repeat regions preventing accurate alignments to the reference sequence and variant calling, which may have led to an underestimation of their genetic diversity, warranting the use of third generation ‘long read’ sequencing technologies and novel approaches to assembling these complex regions. Bioinformatic pipelines that permit detection of these types of variation could be utilised in the future^33,64^. Future studies should take these factors into account, incorporating higher temporal resolution and repeated sampling to better elucidate these dynamics, particularly in the context of parasite antigen diversity and transmission intensity.

In conclusion, we observed differences in the immune responses to malaria antigens, as previously shown ^22,29,65–69^. The variation in antibody responses suggests differential recognition of antigens, likely driven by antibody boosting or memory responses associated with antigen-specific features such as diversity and structural properties. However, the mechanisms underlying the decay of antibodies to these antigens need further investigation, as retrospective analyses indicate variability in antibody waning. The results reinforce that protective antibody responses against clinical malaria are predominantly variant-specific, with repeated exposure to infection necessary to maintain antibody levels and achieve protective immunity to diverse antigens. Taken together, these findings highlight how specific antigen characteristics—including genetic diversity, and structural properties—contribute to differences in immune recognition and protection. Integrating these features into vaccine candidate prioritisation and development pipelines may improve the prediction of vaccine efficacy across diverse populations. Such insights can guide the selection of antigens most likely to generate robust, durable, and broadly protective immune responses. Ultimately, a more detailed understanding of these factors will support the design of next-generation malaria vaccines with improved population-level effectiveness.

## STAR*METHODS

Detailed methods include the following:

- KEY RESOURCES TABLE
- RESOURCE AVAILABILITY

Lead contact Materials availability

Data and code availability

- EXPERIMENTAL MODEL AND SUBJECT DETAILS

Ethical approvals

Study samples: *P. falciparum* patients and negative controls

- METHOD DETAILS

*P. falciparum* antigen selection and expression

Antigen conjugation and Total IgG antibody assay

- QUANTIFICATION AND STATISTICAL ANALYSIS

Data acquisition and normalisation

Multivariate linear regression

Modelling antibody kinetics

Antigen features

Other statistical analyses

## KEY RESOURCES TABLE

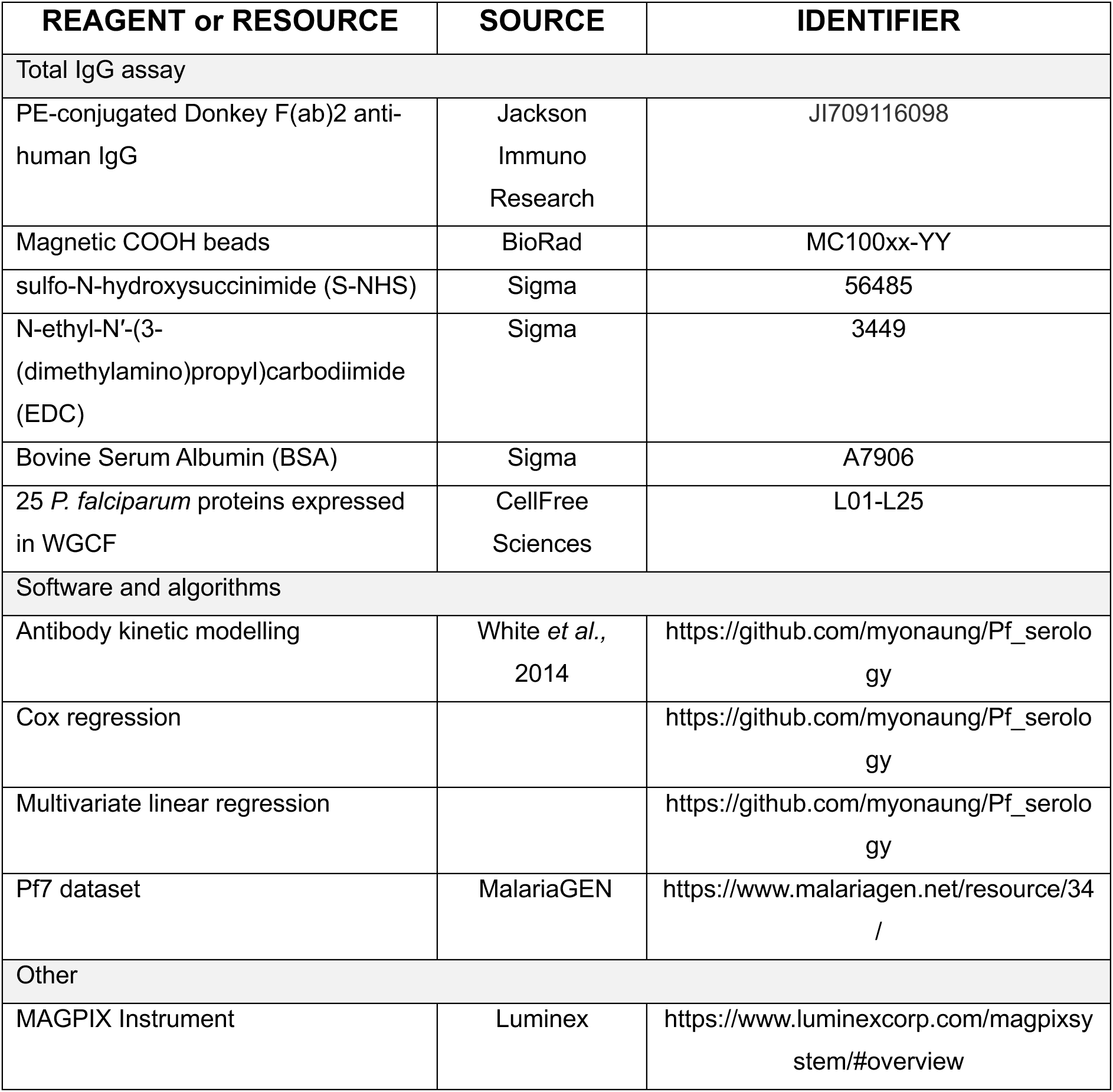

## RESOURCE AVAILABILITY

### Lead contact

Further information and requests for resources and reagents should be direct to and will be fulfilled by the lead contact, Professor Alyssa Barry.

### Materials availability

This study did not generate new unique reagents.

### Data and code availability

- All data are available in the main text or the supplemental information: Data S1.csv contains patient variables from the high transmission cohort and Data S2.csv contains patient variables from the moderate transmission cohort, Data S3.csv contains IgG antibody data (in relative antibody unit) from both cohorts
- The analysis code is available at https://github.com/myonaung/Pf_serology.
- Any additional information required to reanalyze the data reported in this paper is available from the lead contact upon request.

## EXPERIMENTAL MODEL AND SUBJECT DETAILS

### Ethical approvals

The moderate transmission study received ethical clearance from MRAC PNG 09/11, WEHI HREC 09/06, 12/09, Deakin HREC 2023-219. The high transmission cohort study received ethical clearance from MRAC PNG 08/03, WEHI HREC 04/04, Deakin 2023-219. All participants gave informed consent and/or assent to participate in the study. The Human Research Ethics Committee at WEHI (Australia) approved the collection and use of the negative control samples (WEHI HREC 04/04).

### Study samples: *P. falciparum* patients and negative controls

To evaluate antibody dynamics under contrasting exposure conditions—one characterised by frequent boosting due to repeated infections, and the other by minimal boosting in the absence of recurrent exposure—we utilised samples and clinical and parasitological data from two well-characterised paediatric cohorts from the north coast of Papua New Guinea (PNG)^27,28^. These cohorts provided a methodological framework to distinguish the effects of continuous reinfection from those of limited exposure on antibody acquisition and decay. Although the study area experiences year-round endemic transmission, intensified malaria control interventions between 2006 and 2014 led to a substantial reduction in parasite prevalence^46,70,71^, enabling the recruitment of cohorts that capture both high-transmission and moderate-transmission conditions within the same geographical region.

#### High transmission

Plasma samples were obtained from a prospective treatment-reinfection cohort of 206 children aged 5–14 years (median = 9.3) residing in Mugil, Madang Province, PNG, between June and December 2004^28^. On average, each child had 10 *P. falciparum* infection episodes per year, and around one-third (41.2%) of the participants had active blood-stage infections detected by PCR at study enrolment^28,29^. Infections were cleared via artesunate treatment as described^28^. Children were then actively followed every two weeks for symptomatic illness and parasitemia, as well as by passive case detection, over six months. Symptomatic episodes of *P. falciparum* malaria were defined as fever and *P. falciparum* parasitemia >2500 parasites/µl as measured by microscopy. Plasma samples used for this study were those taken at enrolment before treatment with artesunate (n=198), and endpoint of the study (n=156) from a total of 156 children.

#### Moderate transmission

Another cohort study designed as a randomized, double blinded placebo controlled blood- plus liver-stage drug trial was conducted in six villages in the Albinama and Balif areas, Maprik district, East Sepik Province, PNG, between August 2009 and May 2010^25–27^. Each group exhibited a comparable proportion of *P. falciparum* infections following enrolment, with such cases representing 6% of the samples collected within each respective group. This cohort includes 495 children aged 5-10 years (median = 9.6). The risk of *P. falciparum* infection was highly heterogeneous: less than half of the study participants (195 children) experienced a repeated *P. falciparum* infection during the study period (previously exposed individuals), and the average *P. falciparum* infection episode per child per year was less than one^26^. Children were actively followed every two weeks for symptomatic illness and collection of blood samples for PCR detection for the first 12 weeks, and then every two weeks for symptomatic infection and every four weeks for PCR detection. Passive case detection was included over the entire study period. The symptomatic episode of *P. falciparum* malaria was defined as fever and *P. falciparum* parasitemia >500 parasites/µl as measured by microscopy (corrected by qPCR results). Plasma samples used for this study were those taken at enrolment (n=494), before treatment with antimalarials, and at the end point of the study (n=446) with paired samples from a total of 446 children. They included 212 individuals that received blood stage treatment only, and 234 individuals treated with both blood stage and liver stage treatments.

## METHOD DETAILS

### *P. falciparum* antigen selection and expression

A panel of 25 recombinant *P. falciparum* antigens was included in the study. The complete list and details of antigens and their constructs can be found in **Table S1**. Leading vaccine candidate antigens undergoing clinical trials (available at https://clinicaltrials.gov/expert-search?term=malaria%20vaccine), antigens known or predicted to be involved in erythrocyte invasion, and antigens under immune selection were included ^7,22,29,31,72^. All antigens were expressed using a wheat germ cell-free (WGCF) system (CellFree Sciences, Matsuyama, Japan). All antigens were expressed and purified as previously described in detail ^63,73,74^. Two distinct constructs of PF3D7_0731500 (erythrocyte binding antigen-175, EBA175 RII and EBA175 RIII-V), each representing a different region of the protein or sequence from the 3D7 strain, were included, bringing the total to 25 antigens. Pfs25, given its exclusive role in the vector stage development, was used as a negative control.

### Antigen conjugation and Total IgG antibody assay

The conjugation of purified *P. falciparum* antigens to magnetic COOH microspheres (Bio-Rad Laboratories, Inc., California, USA) was performed as described ^63,74^. The concentration of each antigen used for conjugation was optimized to achieve a standard curve with log-linearity over the range of 0.04–8 µg (data available at https://github.com/myonaung/Pf_serology/blob/main/protein_construct_metadata.xls x). In brief, 2.5 × 10^6^ microspheres with unique fluorescent colours were activated before the optimal concentration of an antigen was added and incubated for two hours at room temperature in the dark before excess antigens were removed.

### Total IgG data acquisition and normalisation

Total IgG antibody measurements were performed using a multiplexed bead-based assay as previously described ^63,74^. Briefly, 500 antigen-conjugated microspheres per well were added into a 96 well-plate and incubated at room temperature with plasma samples diluted in a 1:100 ratio. Antigen-coupled microspheres were then incubated with a PE-conjugated anti-human secondary antibody at 1:100 dilution (donkey F (Ab’)2 anti-human IgG Fc, 1 mg/ml, Jackson ImmunoResearch Laboratories, Inc., catalogue number JIR709116098). Total IgG levels were measured at the longitudinal cohorts’ first and last time points. A standard curve obtained from positive control plasma with 2-fold serially dilution (ranging from 1:50 to 1:51200) was included on every plate. This positive control plasma was obtained from hyper-immune individuals in PNG ^63,74^. Plasma from Australian Red Cross malaria-naïve individuals with no known previous exposure to malaria (n = 25) were used as negative controls. The assays were performed on a Luminex® 200™ (Luminex® Corporation, Texas, USA) platform.

Median fluorescence intensity (MFI) values were converted into arbitrary relative antibody units (RAU) based on the antigen-specific standard curves on each plate to adjust for any plate-plate variations. Raw data are available at Data S1-S3. The conversion was performed using a 5-parameter logistic regression model ^63,74^. Seropositivity is defined based on Melbourne Red Cross malaria naïve samples (n = 25). We use a threshold of mean RAU plus three standard deviations from naïve sera. RAU of each antigen above this threshold are defined seropositive. The seroprevalence for an antigen is calculated as follows:

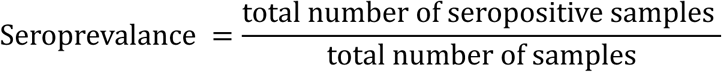

### Multivariate linear regression

To measure the association between antibody levels and the risk of symptomatic *P. falciparum* malaria, we used *Cox regression* analysis based on the time to the first episode of clinical or symptomatic malaria (defined as concurrent parasitemia ≥2500 parasites/µL for the high transmission cohort or ≥500 parasites/µL for the moderate transmission cohort with axillary temperature ≥37.5°C or history of fever in preceding 48 hr). MFI values were interpolated into RAU based on the parameters estimated from the plate’s standard curve as previously described^75^. The observed antibody levels were then stratified into tertiles for each transmission setting and transformed into 0, 1, and 2 for the low, medium, and high antibody tertiles, based on the level of sero-reactivity to each of the 25 antigens included in the study. We fitted adjusted *Cox regression* models for both cohorts adjusting for known confounders ^22,26,28,29^ – log10-transformed age, locations, blood-stage *P. falciparum* detection at the first timepoint, *P. falciparum* parasitemia count (parasite per µl) and infection with other *Plasmodium* species. Adjusted hazard ratios (aHR) were calculated as previously^22,26,28,29^. In the high-transmission cohort, we report aHR comparing medium versus low antibody tertiles, while in the moderate-transmission cohort, we report high versus low tertiles; these contrasts provided the largest effective sample size within each setting and therefore yielded the most precise and stable estimates. Adjusted Hazard ratio (aHR) values below 1 are associated with protection. *Potential Protective Efficacy* (PPE) was calculated as [1-aHR]*100 as described previously ^29^. We used *survival* R package.

To identify factors driving variation in antibody responses among individuals at the endpoint of the study, multivariate linear regression models were used. This retrospective analysis was conducted using antibody responses of each antigen (in *RAU*) from the last follow-up time point as a reference. Initial analysis revealed strong collinearity between the number of detected infections and time since last infection (r = -0.573, p < 0.001), which compromises the reliability of relative importance estimates when both continuous variables are included simultaneously. To address this methodological challenge, we implemented categorical modelling approaches alongside the original continuous variable models and excluded variable pairs with moderate to strong correlation (|r| > 0.6), ensuring that only one variable from each correlated pair was included in the model. Additionally, original model without transformed variables and ridge regression were employed as alternative approaches and the best model was chosen based on total R^2^ explained variance with the higher value, the better the model. The final model included: (i) log10-transformed age (serving as a proxy for lifetime exposure); (ii) infection timing categories (Very Recent ≤30 days, Recent 31-90 days, Moderate 91-180 days, Distant >180 days since last *P. falciparum* infection); (iii) *P. falciparum* parasitemia categories *(None, Low ≤500, Medium 501-5000, High >5000 copies/μL)*; (iv) the number of detected *P. vivax* infections during the follow-up period and (vi) an error term capturing any additional unexplained variation. The total variance of each variable was normalised to 1 and was the sum of variance given by both explained and unexplained variance, respectively. The contribution of each factor to the total variance was estimated using the *relaimpo* and *car* R packages for standard linear models. The script is available at https://github.com/myonaung/Pf_serology.

### Modelling antibody kinetics

A previously validated mathematical model^34^ was adapted to retrospectively understand the antigen-specific antibody kinetics in a cross-sectional framework based on the measured antibody data of different antigens from the last timepoint^34,76^. This Bayesian non-linear mixed effects model captures the complex dynamics of antibody responses following *P. falciparum* infection by modelling antibody levels as a function of time since last reported infection. Given the high frequency of repeated infections in the high-transmission cohort, which complicates accurate timing of infection events, we restricted our analysis to the moderate-transmission cohort to better align with the model’s assumptions and improve interpretability of antibody decay patterns. Similar to previously published model^34^, it incorporates biphasic decay patterns and contributions from distinct antibody-secreting cell populations with different longevity profiles—short-lived, intermediate-lived, and long-lived antibody secreting cells (ASCs)—with the key novel addition of strain-specific variation, based on the known haplotype prevalence of the 3D7 allele. The antibody response follows a mechanistic framework where observed antibody levels are modeled as:

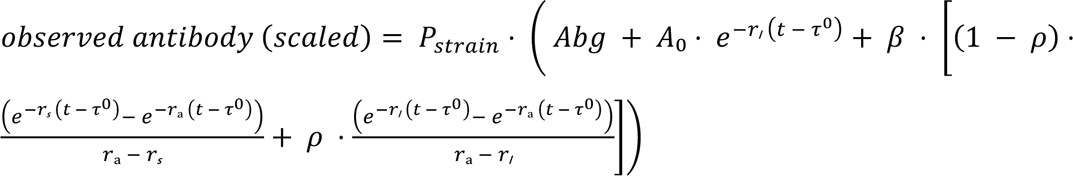

where t represents scaled time since last *P. falciparum* infection. The model parameters capture distinct biological processes: *Abg* represents background antibody levels reflecting baseline immunity maintained from cumulative infection history; *A_0_* quantifies the antibody level at the time of last infection; *β* represents the amplitude of a complex decay component that captures population-level variation in antibody kinetics; *r_l_, r_s_*, and *r_a_* are decay rates for long-lived, short-lived, and intermediate-lived ASCs estimated at the population level, respectively; *ρ* represents the proportion of ASCs that are long-lived (0 ≤ ρ ≤ 1); *τ*_0_ is a temporal shift parameter that accounts for uncertainty in reported infection dates and population variation in peak antibody response timing; and *P_strain_* is a strain-specific probability factor based on 3D7 haplotype prevalence of each antigen in the PNG population based on Pf7 database.

The hierarchical model structure allows *Abg* to vary by infection history categories (No Infections, 1-2 Infections, >2 Infections) while incorporating random effects for individual participants to account for inter-individual variation. Initial antibody boost (*A_0_*) and scaling factor (*β*) vary only by infection category, while decay parameters (*r_l_, r_s_*, *r_a_*), proportion parameter (*ρ*), and temporal shift (*τ_0_*) remain fixed across all participants. This mixed-effect model accounts for inter-individual variation by estimating random effects for each participant, while also capturing cohort-level trends across infection history groups. Both antibody levels and time since infection were scaled to improve model convergence and parameter interpretation. Bayesian inference was implemented using the *brms* R package with biologically relevant prior information, with exponential distributions for decay rates and normal or beta distributions for other parameters. The same prior distributions were applied consistently across all antigens to ensure comparability of parameter estimates. The student-t distribution was chosen as the likelihood function to robustly handle potential outliers and heavy-tailed distributions in antibody measurements. Detailed modelling parameters are available on https://github.com/myonaung/Pf_serology. Model performance was evaluated using posterior predictive checks to assess coverage rates of 95% prediction intervals and Bayesian R² to quantify predictive accuracy. Coverage rate improvement measures how well strain-specific models capture observed data within prediction intervals compared to strain-agnostic models, while mean accuracy improvement quantifies the difference in explained variance between model formulations. These metrics provide complementary assessments of model predictive performance, with positive improvements indicating better performance of strain-specific modelling approaches. The model quantifies antibody half-lives for each ASC population and estimates the relative contribution of long-lived versus short-lived responses in a strain-specific context. However, due to data limitations, we did not calculate precise half-life values for each antigen. Instead, we computed a composite normalized antibody durability score that integrates modelled half-life estimates from three response components—short-lived, intermediate, and long-lived—using a weighted average (20%, 20%, and 60%, respectively). Antigens with the longest derived half-life estimates received a normalized antibody durability score of 1, whereas those with the shortest derived half-life estimates received a score of 0. The resulting composite scores provide a relative, population-level index of antibody persistence across antigens.

### Source of whole genome sequences (WGS), variant calling and antigen features

Characteristics of each antigen were determined to identify features associated with antibody responses. To measure population level diversity, we utilised whole genome sequences (WGS, n= 222 *P. falciparum* isolates from Papua New Guinea) ^34,35^ included by the Malaria Genomic Epidemiology Network (MalariaGEN) Pf7 project release^79^. The raw WGS data downloaded from the European Nucleotide Archive was processed by mapping high quality reads to the 3D7 reference genome (PlasmoDB v68) using *GATK v4.1.4.0* as mentioned previously^5,79^. We restricted our analyses to single nucleotide polymorphisms (SNPs) from 14 nuclear chromosomes only excluding indels and variants from hypervariable regions. Further hard filtering was applied based on the mapping quality and genotype missingness (10% of total samples) to obtain high-quality SNPs as mentioned previously^5^. Singleton SNPs (i.e. polymorphisms seen only once in the entire dataset) were assumed to be artefacts and converted back to reference to prevent false positive variants. No minor allele frequency threshold was applied to the dataset, and we limited our analyses to major clone.

Polymorphisms (SNP) data from PNG (n = 222) was used to estimate the genetic diversity of each vaccine candidate antigen based on expressed regions included in this study. *Tajima’s D* (*D*)^80^ was calculated with a sliding window approach (50 base pair window with step size of 5) for each antigen. To focus on balancing (immune) selection for each antigen, the analysis was restricted to positive values, excluding SNPs with a negative *D* value. We then assigned a weighted average *D* or *D’* value for each antigen based on SNPs with values above values. These calculated *D’* scores were combined to reflect the degree of balancing selection per antigen, providing a reasonable estimate of immune selection, though more precise methods, such as identifying immune selection hotspots within each antigen, may yield even finer insights^52^. Haplotype diversity was calculated as previously described^5^.

Each antigen, based on the 3D7 reference, was also annotated with amino acid secondary structure (*ragp* R package), and intrinsically disordered properties (*IUPred3*). All analyses were performed using default parameters.

### Other statistical analyses

To compare relative antibody units between the first timepoint and endpoint for each antigen within each cohort, as well as diversity and physicochemical metrics between defined groups, we performed the *Mann-Whitney U test* (*Wilcoxon rank-sum test*) using *wilcox.test()* function from *stat* R package.

## Supplemental information

Document S1. Figures S1–S3, Tables S1 and S2, and supplemental references

## Data Availability

All data produced in the present work are contained in the manuscript.

## Acknowledgements

We thank all the study participants and their families in PNG. We acknowledge the efforts of the original study teams for collecting the *Plasmodium* clinical samples. We also thank Dr. Balu Balan (WEHI) for assistance with intrinsic disordered region prediction. This work was supported by the National Health and Medical Research Council (NHMRC) of Australia (Investigator Grant GNT1173210 to RJL, Project Grant 1161066 to AEB, Synergy Grant 2018654 to IM and AEB). IM, AEB and RJL are members of the Australian Centre for Research Excellence in Malaria Elimination, funded by the NHMRC (GNT2024622). The authors acknowledge the Victorian State Government Operational Infrastructure Support and Australian Government NHMRC IRIISS. The funders had no role in study design, data collection and analysis, decision to publish, or preparation of the manuscript.

## Inclusion and Diversity

The author list of this paper includes contributors from the location where research was conducted who participated in the data collection, design, analysis, and/or interpretation of the work.

