## Supplementary material for "Protective immunity to clinical malaria is modified by the genetic diversity of *P. falciparum* antigens": Document S1.pdf

**Table S1: Plasma samples included in the study**

| Cohort | Study timepoint | n |
|---|---|---|
| Moderate transmission | Enrolment | 494 |
| | Endpoint | 446 |
| Total | Matched enrolment + endpoint samples from same individuals | 446 |
| High transmission | Enrolment | 198 |
| | Endpoint | 156 |
| Total | Matched enrolment + endpoint samples from same individuals | 156 |

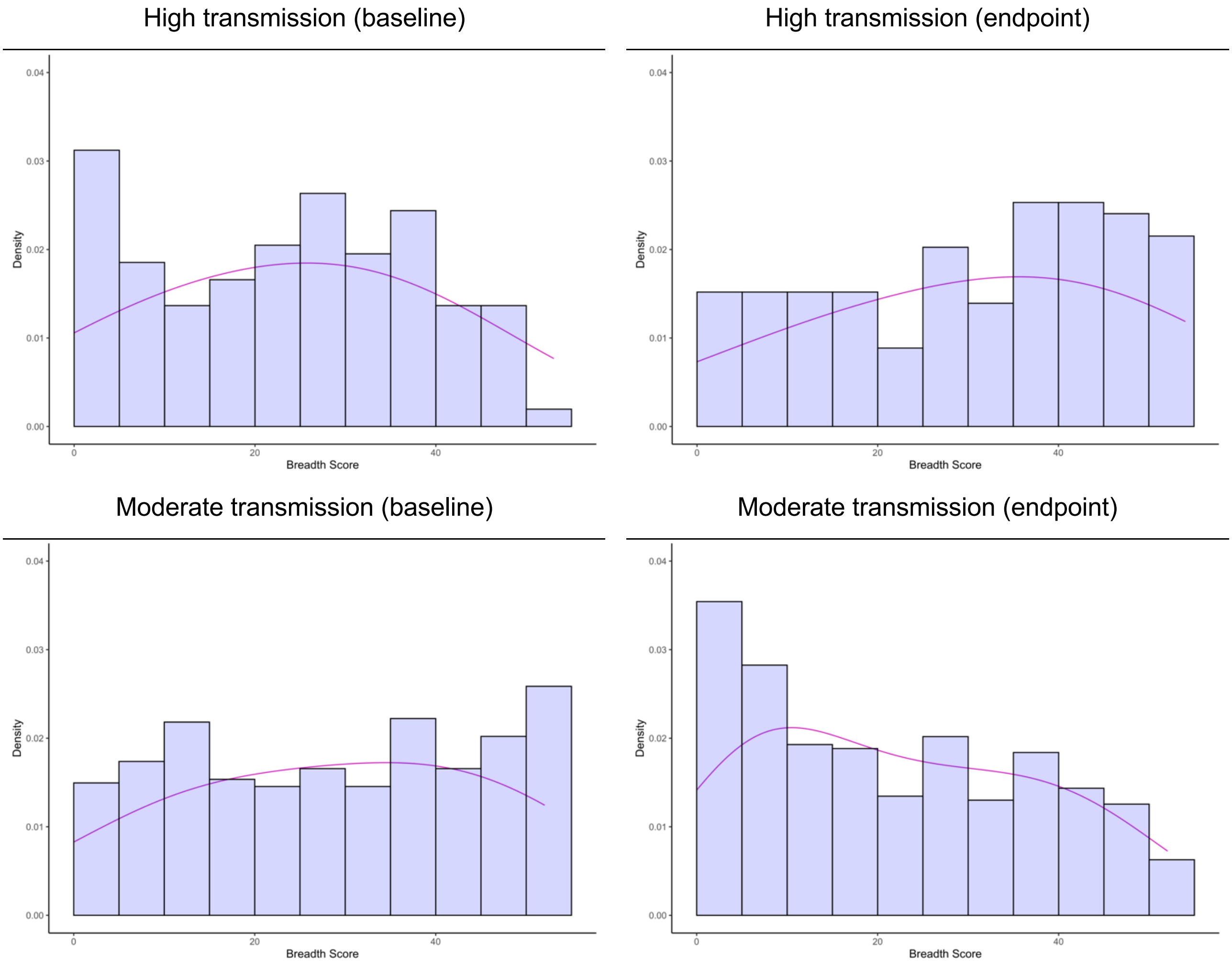


**Figure S1.** Breadth of anti-*P. falciparum* IgG antibodies in young PNG children at different transmission settings. To quantify antibody breadth, IgG responses to 25 *P. falciparum* antigens were stratified into tertiles for each antigen and assigned scores of 0, 1, or 2 corresponding to low, medium, and high antibody levels, respectively. Individual scores were then summed across all antigens to generate a composite breadth score

A. Moderate transmission cohort

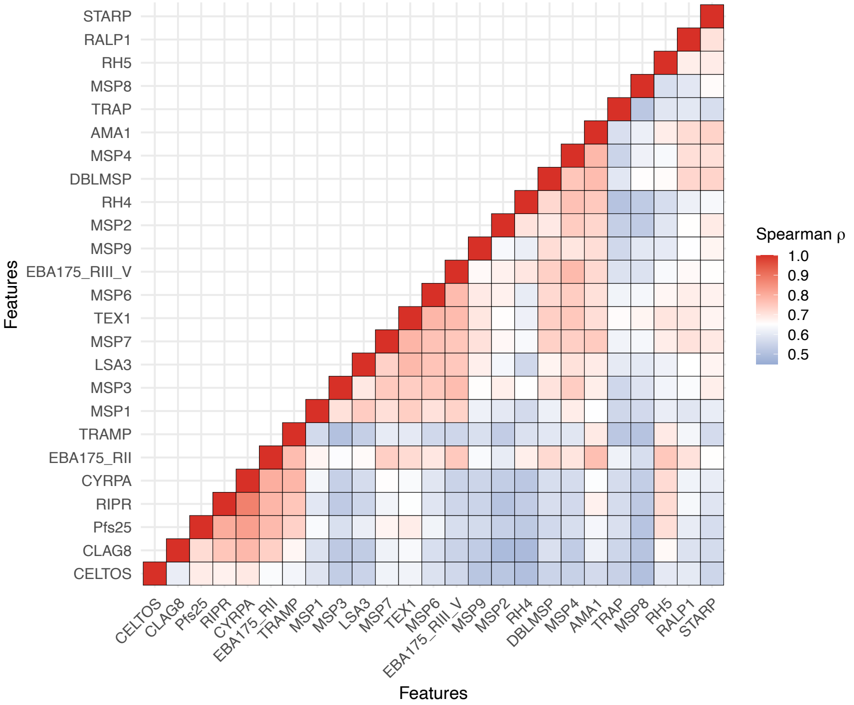


B. High transmission cohort

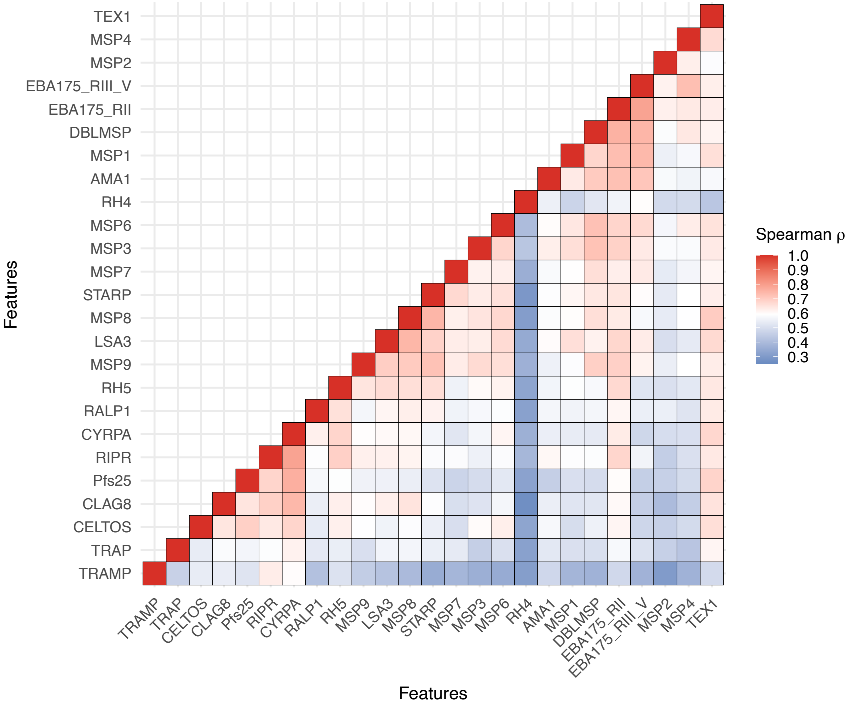


**Figure S2.** Correlations between IgG to *P. falciparum* vaccine candidate antigens in different transmission settings. Correlation coefficients between antibody levels to every pair of antigens were calculated using Spearman's rank correlation tests.

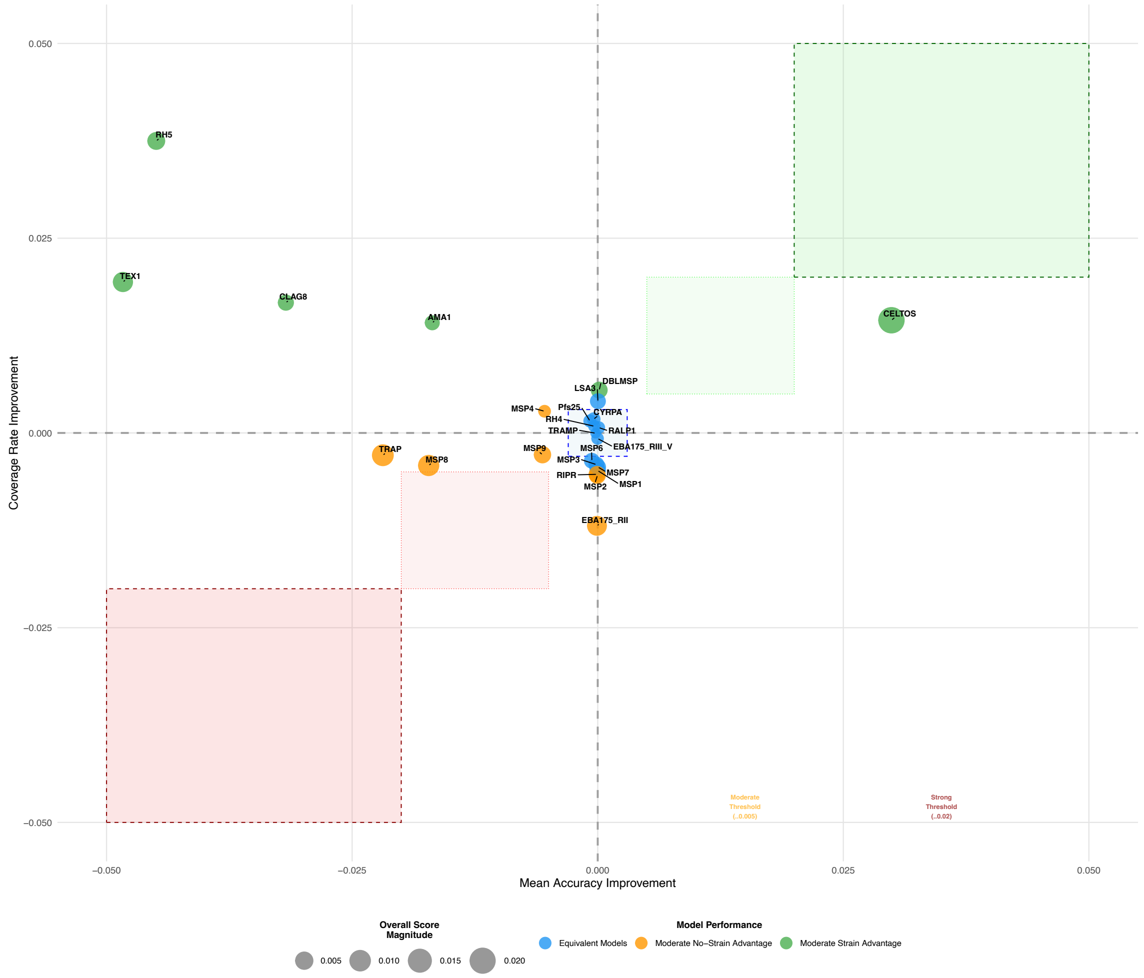


**Figure S3.** Comparative performance of strain-specific vs. strain-agnostic antibody kinetics models across 24 antigens at moderate transmission settings. Each point represents an antigen, positioned according to its improvement in 95% prediction interval coverage (y-axis) and mean predictive accuracy (Bayesian $R^2$; x-axis) when using a strain-specific model compared to a strain-agnostic counterpart. Points are colored by overall model performance categories based on threshold analysis: green indicates strain-specific advantages (moderate to strong), blue represents equivalent model performance, and orange indicate no-strain model advantages. Point size represents the Overall Score Magnitude, with larger points indicating stronger performance differences between the two modelling approaches. The dotted lines show classification thresholds (±0.005 for moderate, ±0.02 for strong differences). Points in the upper-left quadrant indicate antigens where strain-specific models produce better-calibrated uncertainty estimates despite slightly reduced point-prediction accuracy, potentially reflecting improved generalization. Upper-right quadrant points suggest antigens where strain-specific models offer improvements in both calibration and accuracy. Analysis excludes STARP (extreme outlier) to focus on the main cluster of antigens. Strain-specific models were prioritized when robust uncertainty quantification and model interpretability were emphasized over pure point-prediction accuracy.

**Table S2:** Descriptions of antigens included in the study

| Life cycle stage | Antigen | AA length | Expression region |
|---|---|---|---|
| Pre-erythrocytic | LSA3 | 1559 | aa:750-1433 |
| | STARP | 595 | Full length |
| | TRAP | 575 | Full length |
| | CelTOS | 183 | Full length |
| Erythrocytic | AMA1 | 623 | aa:25-546 |
| | CLAG8 | 1394 | aa:707-1391 |
| | CyRPA | 362 | aa:27-362 |
| | DBLMSP | 698 | aa:26-697 |
| | EBA175 | 1503 | Region II (aa:142-764) |
| | | | Region III-V (aa:761-1298) |
| | MSP1 | 1721 | aa:1607-1699 |
| | MSP2 | 273 | aa:26-249 |
| | MSP3 | 355 | aa:27-354 |
| | MSP4 | 273 | Full length |
| | MSP6 | 372 | aa:23-371 |
| | MSP7 | 352 | aa:24-351 |
| | MSP8 | 598 | aa:1-597 |
| | MSP9 | 744 | aa:1-743 |
| | RALP1 | 750 | aa:396-710 |
| | RH4 | 1717 | aa:28-766 |
| | RH5 | 527 | aa:26-526 |
| | RIPR | 1087 | aa:279-995 |
| | TEX1 | 1104 | Full length |
| | TRAMP | 2266 | aa:26-309 |
| Gametocyte | Pfs25 | 218 | aa:16-194 |
